# Use of face coverings in public during the COVID-19 pandemic: an observational study

**DOI:** 10.1101/2020.06.09.20126946

**Authors:** Nicholas L. Arp, Tung H. Nguyen, Emma J. Graham Linck, Austin K. Feeney, Jonathan H. Schrope, Katrina L. Ruedinger, Anqi Gao, Margot Miranda-Katz, Ashley E. Kates, Nasia Safdar

## Abstract

Public health agencies have recommended that the public wear face coverings, including face masks, to mitigate COVID-19 transmission. However, the extent to which the public has adopted this recommendation is unknown. An observational study of 3,271 members of the public in May and June 2020 examined face covering use at grocery stores across Wisconsin. We found that only 41.2% used face coverings. Individuals who appeared to be female or older adults had higher odds of using face coverings. Additionally, location-specific variables such as expensiveness of store, county-level population and county-level COVID-19 case prevalence were associated with increased odds of using face coverings. To our knowledge, this is the first direct observational study examining face covering behavior by the public in the U.S., and our findings have implications for public health agencies during the COVID-19 pandemic.

## Introduction

The Centers for Disease Control and Prevention (CDC) recommends that the public wear face coverings as a major non-pharmaceutical intervention to mitigate COVID-19 transmission^1^, particularly when physical distancing is difficult. Since the United States does not have a culture of face covering use by the public and there have been reports of violent retaliation by people asked to wear a face covering^2^, there is uncertainty regarding the extent to which this recommendation has been adopted. A recent survey by Gallup reported 68% of U.S. adults claim to ‘always’ or ‘sometimes’ wear a face covering in public.^3^ To date, however, there have been no direct observational studies examining face covering usage by the general public in the United States. The objective of this study was to quantify face covering usage by the public visiting grocery stores using a convenience sample of Wisconsin residents.

## Methods

We used direct observations of individuals exiting 26 grocery stores to assess face covering use across 20 counties in Wisconsin between 16 May and 1 June 2020 (Figure 1A). We chose to observe face covering usage at grocery stores because they provide essential services, are visited frequently by the public, and present settings where reliable physical distancing may be challenging. No stores we observed required face coverings upon entry. The stores were selected based on geographic convenience for the observers. The time of the start of observations, through a retrospective analysis of all observations, were normally distributed with a mean of 2:45PM and standard deviation of 105 minutes. Each observer recorded the shoppers’ apparent age (minor, young adult, adult, older adult), gender expression (female/male), and face covering use (present/absent: any type of cloth covering, surgical face mask, or N95 respirator). Inter-rater reliability was assessed for 307 observations from two simultaneous observers using Cohen’s kappa coefficients.

**Figure 1.**
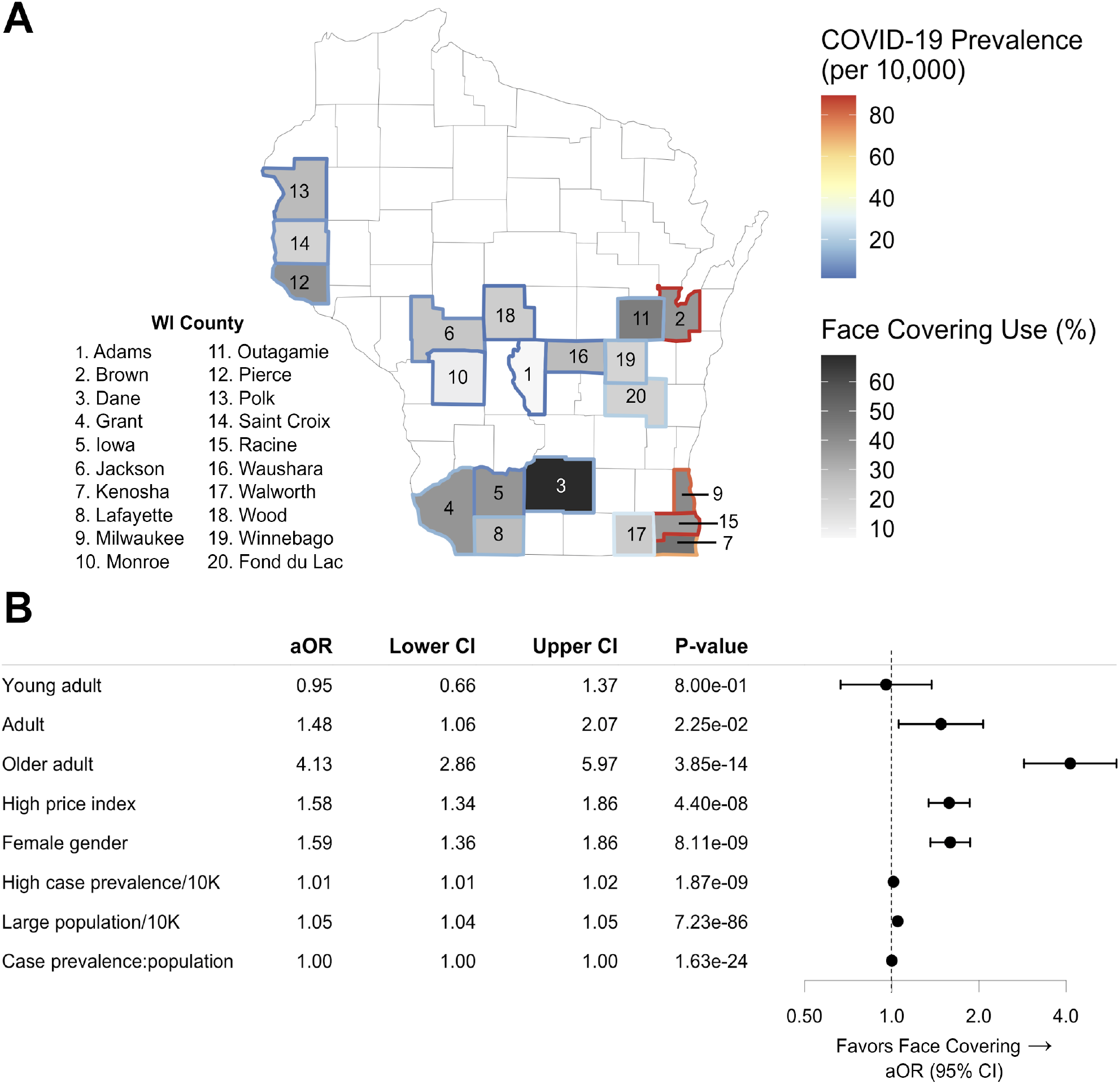
**A**. Map of Wisconsin counties represents observation locations where face covering use was quantified. Color of county outline indicates case prevalence per ten-thousand cases. Fill shade intensity represents the percentage of total individuals that wore a face covering. **B**. Adjusted odds ratios (aOR) and 95% confidence interval (CI) of face covering usage were calculated and plotted from multiple logistic regression. The aOR for age is in reference to the odds of face covering use by minors. All variables included in the model are shown in the table.

The price index for each store was calculated as a relative z-score based on the price of 12 staple food items — onion, potato, apple, soda, yogurt, milk, the least and most expensive dozen large eggs, the least and most expensive chicken breast, and the least and most expensive butter at standard units — to determine if face covering use was associated with store expense.

We used multiple logistic regression to examine associations between age category (in reference to minors), gender expression, price indices, total county population, and county-level COVID-19 case prevalence^4^ on mask usage. Standardization of population was done to match the units of case prevalence (i.e. 10,000 becomes 1, 100,000 becomes 10). In the same model, we also assessed the interaction between COVID-19 case prevalence and total county population on mask usage. Adjusted odds ratios (aOR), including these covariates, 95% confidence intervals, and Wald test p-values were calculated. Analyses were conducted using glm in R version 4.0.

To determine the representativeness of the sample, we used a two-sided Kolmogorov-Smirnov (KS) test to evaluate whether the U.S. Census Tract of the observed locations reflected the distribution of race (percent non-white) and median family income reported across Wisconsin.^5^

## Results

We observed a total of 3,271 individuals, 41.2% of whom were observed wearing face coverings when exiting grocery stores. There was a higher prevalence of face covering use by older adults (59.5%) compared to minors (26.2%), young adults (34.8%), and adults (39.9%); and by females (44.8%) compared to males (36.9%) (Table 1).

**Table 1.**
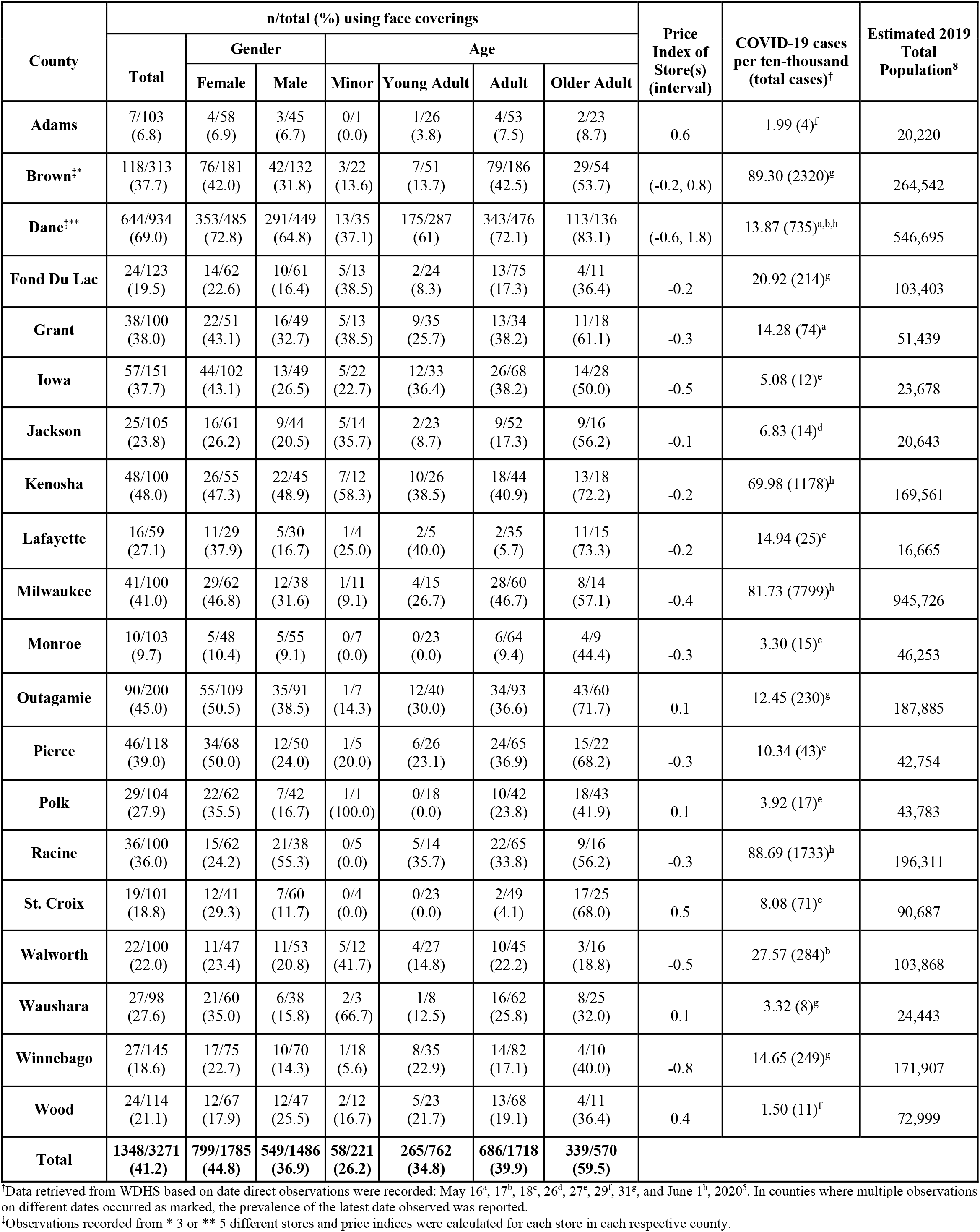
Multi-county observation data of face covering use by the public at grocery stores in Wisconsin.

In multiple logistic regression analysis, we found that age categories of adult (aOR = 1.48, 95% CI = 1.06-2.07, p-value = 2.25e-02) and older adult (aOR = 4.13, 95% CI = 2.86-5.97, p-value = 3.85e-14), female gender (aOR = 1.59, 95% CI = 1.36-1.86, p-value = 8.11e-09), and observations at higher price index stores (aOR = 1.58, 95% CI = 1.34-1.86, p-value = 4.40e-08) were statistically significantly associated with higher odds of face covering usage (Figure 1B). In addition, case prevalence (standardized to cases per ten thousand) was moderately associated with face covering usage (aOR = 1.01, 95% CI = 1.01-1.02, p-value = 1.87e-9). Total population, converted to population per ten thousand, was moderately associated with face covering usage (aOR = 1.05, 95% CI = 1.04-1.05, p-value = 7.23e-86). Although case prevalence and population were positively associated with face covering usage, the significant interaction between case prevalence and population suggests heterogeneity in these effects.

To explore this heterogeneity further, we subset the county-level observations seen in Table 1 to the top five most populous counties (Milwaukee, Dane, Brown, Kenosha, and Racine), and we subsequently observed an inverse linear association between case prevalence per ten thousand and percentage of face covering usage (slope = -0.42% per unit increase in case-prevalence, Pearson’s r = -0.99). All Dane county observations took place in Wisconsin’s capital, Madison, which was an outlier with a high percentage of face covering usage and low case prevalence. However, this negative correlation was not observed with the remaining counties outside the five most populous counties (analysis not shown).

The Cohen’s kappa coefficients for age (0.79, ‘substantial agreement’), gender expression (0.98, ‘almost perfect agreement’), and face covering usage (0.92, ‘almost perfect agreement’) indicate these variables were robustly collected across observers independently. Additionally, we found no significant difference between the convenience sample and Wisconsin at large using the KS test (median income: p = 0.751, D = 0.145; percent non-white: p = 0.203, D = 0.24).

## Discussion

During May and June 2020, the United States was in the midst of the COVID-19 pandemic with many states, including Wisconsin, initiating plans for reopening after months of stay-at-home orders. However, during this time period, we found face covering usage in public was not widely practiced, despite recommendations by multiple public health agencies, including the Wisconsin Department of Health Services.^4^ A previous study modeled that 80% compliance of face covering use by the public demonstrated the greatest decrease in disease burden and COVID-19 transmission.^6^ Our findings report that the Wisconsin public is achieving approximately half this rate of compliance. It is reassuring to report that there was a higher prevalence of face covering use by older adults as these individuals are at higher risk for the severe complications of COVID-19.^7^

Our study has limitations due to its cross-sectional design, use of convenience sampling and its lack of observations in northern Wisconsin. Face covering misclassification could occur if the face covering was removed prior to the observation at the store’s exit door. Further, gender expression and apparent age could have been misclassified due to observer bias. However, reassuringly, inter-rater reliability was determined to be high for these observed variables.

Our results have important implications for public health agencies. Our results suggest the need to develop and test interventions to promote face covering usage by the general public in the United States. Future directions from this report include examining the reasons why some individuals choose not to wear face coverings in public.

## Data Availability

All data is available from the corresponding author upon request.

